# Somatic Mutation Profiles in Colorectal Cancers Differ by Population

**DOI:** 10.64898/2026.06.24.26356431

**Authors:** Batsirai M. Mabvakure, Patharapa Promprasert, Lucia Martinez Cruz, Siddhi Patil, John Barros, Matthew Hayhurst, Elham Mohebbi, Deborah de la Caridad Delgado Herrera, Grace Lee, Sophia Latif, Fiona Williams, Rashmi Samdani, Anju Duttargi, Bereket Berhane, Esmael Besufikad, Selamawit Tadesse, Aisha Jibril Suleiman, Christina Lefante, Mei-Chin Hsieh, Kristen Purrington, Ernest Adjei, Tingting Qin, Maureen Sartor, Elena M. Stoffel, Laura S. Rozek

**Author notes:** **Corresponding author:** Laura Rozek, Professor of Oncology, Georgetown University.

## Abstract

**PURPOSE:** Colorectal cancer (CRC) incidence and mortality rates differ by population, and evidence suggests that genetic differences may affect cancer biology. However, studies investigating CRC variants in genetically heterogeneous populations are limited. Using somatic tumor mutation profiling of CRCs diagnosed in African Americans (AAs), Ghanaians, Ethiopians, and NHWs, we explore correlations between population group and population-specific tumor variants.

**PATIENTS AND METHODS:** Somatic DNA from CRC tumors resected from 150 individuals, including 43 AAs (27%), 53 NHWs (35%), 21 Ghanaians (14.2%), and 33 Ethiopians (22.3%), was sequenced on the Illumina NovaSeq platform, targeting 290 genes. We compared mutations in AAs, Ghanaians, and Ethiopians to those in NHWs to identify variants enriched in historically underrepresented groups.

**RESULTS:** US cohort tumors were diagnosed at significantly younger ages with more early-onset cases (<50 years old) than African cohorts (*p* <0.05). Significant differences were observed in primary tumor location, MMR phenotypes, *KRAS* mutations, and distribution of tumor mutational burden by population. *BRAF V600E* mutations were rare across all groups, while non-*V600E BRAF* mutation rates were higher in AA and NHW (43-44%) than Ethiopian and Ghanaian (14-33%) samples. Population-specific differences were identified in mutation rates of *APC, CTNNB1, RNF43, PIK3CA,* and *TP53*, as well as in pathogenic variant occurrence.

**CONCLUSION:** Geographic origin and age of onset appear to shape the CRC mutational landscape. The similarity between AA and NHW tumor profiles likely reflects their shared predominance of early-onset cases, suggesting that age- and geography-associated factors, including environmental exposures, may influence tumor phenotypes.

## INTRODUCTION

The comprehensive molecular characterization of colorectal cancer (CRC) by the Cancer Genome Atlas Network (TCGA) revealed significant insights into the variability of the genetic landscape and associated clinical relevance (1,2). The TCGA represented an important first step in our understanding of colorectal carcinogenesis; however, 297/374 (79%) of CRC tumors in the population were from individuals of northern European descent (3,4). African Americans (AAs) comprised fewer than 20% of study subjects (5). Differences in tumor somatic mutation profile may explain at least part of the documented racial disparities in cancer prognosis, as has been observed for lung, prostate, and breast cancers (6–10). For example, triple-negative breast cancers (TNBC) is associated with poor prognosis and is more likely to be diagnosed in individuals of African ancestry with specific African-associated tumor immunogenic profiles (11,12).

We hypothesize that studies that include populations across the African diaspora will identify novel pathways associated with the aggressiveness of the tumors. Additionally, similar to CRC among AAs, CRC incidence is increasing among Africans, with African CRC patients presenting at younger ages and with more advanced tumors when compared to North American CRC patients (13–15). Although studies have been conducted in African-ancestry populations (16–18), there remains a critical gap in understanding the role of tumor somatic mutation phenotypes and how they influence CRC pathogenesis among different population groups. A few recent efforts drawing on TCGA, GENIE, and large clinico-genomic datasets have also identified ancestry-associated differences in somatic drivers and coalteration patterns, but these cohorts include very few patients from contemporary African populations and almost no side-by-side comparisons of African Americans with East and West Africans (15,17). The purpose of this study was to examine somatic mutation profiles of tumors collected from AAs, non-Hispanic White Americans (NHW), Ethiopians, and Ghanaians and to identify patterns of genomic variation in CRC among these populations that may impact tumor biology and response to treatment.

## MATERIALS AND METHODS

### Study population, ethical approval, and collecting samples

We collected and analyzed archived formalin-fixed paraffin-embedded (FFPE) CRC tumors previously resected from individuals undergoing clinical care in Komfo Anoyke Teaching Hospital (KATH), Kumasi, Ghana; St Paul’s Millenium Medical College, Addis Ababa, Ethiopia; and the University of Michigan and Georgetown University in the US. Demographic information (age and sex) and clinical information (tumor stage, differentiation, and primary tumor location) were abstracted for all cases. Early-onset CRC was defined as diagnosis before age 50, and sampling was intentionally performed to overrepresent early-onset cases.

### Immunohistochemical staining for mismatch repair proteins

Tumor slides were reviewed by clinical pathologists at Georgetown University and KATH, and areas of high tumor cellularity were identified for microdissection using 5–10 unstained 5 µm sections. Hematoxylin and eosin (H&E)-stained slides were used to estimate tumor and normal cell content. Immunohistochemical (IHC) staining for mismatch repair (MMR) proteins *MLH1*, *PMS2*, *MSH2*, and *MSH6* was performed on FFPE tissue sections using the Dako Autostainer Link 48 platform. Sections (4 µm) underwent deparaffinization, rehydration, and heat-induced epitope retrieval (HIER) using Envision FLEX Target Retrieval Solution at high or low pH, depending on the antibody, followed by peroxidase blocking, primary antibody incubation, HRP secondary detection, DAB chromogen development, and hematoxylin counterstaining. Antibodies were sourced from Cell Marque: *MLH1* (Clone G68-728, 1:25), *PMS2* (Clone MRQ-28, 1:100), *MSH2* (Clone Y69, 1:300), and *MSH6* (Clone 44, 1:50). Loss of MMR protein expression was defined as absence of nuclear staining in tumor cells with preserved expression in adjacent non-neoplastic cells.

### Nucleic acid extraction

Tumor DNA was extracted from the FFPE sections with ≥60% of tumor cellularity using the QIAGEN QIAamp FFPE tissue kit (Catalog Number: 56404) on a QIAcube Connect automated platform. Samples were deparaffinized and treated with Proteinase K prior to extraction. DNA concentration and purity were assessed by Nanodrop (260/280 and 260/230 ratios). Samples with DNA concentrations > 30 ng/μL and 260/230 ratios of 2.0 – 2.2 were selected for sequencing.

### Library Preparation and targeted DNA Illumina sequencing

Targeted DNA sequencing was performed using the QIAseq Targeted DNA Pro Human Comprehensive Cancer Research Panel (Qiagen, Cat. No. 333665; Panel ID: PHS-3000Z), which targets 290 cancer-associated genes using hybrid-capture–based enrichment (**Table S1**). Libraries were prepared from 500 ng of genomic DNA incorporating unique molecular indices (UMIs) for error correction. Library quality was assessed using the Agilent Bioanalyzer; libraries with average fragment sizes of 180–250 bp and concentration ≥30 ng/µL were used for sequencing. Sequencing was performed on the Illumina NovaSeq platform at the University of Michigan Genomics Core, targeting ≥6 million reads per sample at ×1000 depth. The panel enables detection of SNVs, indels, and CNVs with high sensitivity.

### Bioinformatics analysis

Sequencing quality was assessed using FastQC v0.11.9 and samtools, and results were aggregated with MultiQC (v1.28). Raw FASTQ files were processed in QIAGEN GeneGlobe for UMI-based error correction, alignment to GRCh38, variant calling, and annotation. Additional variant annotation was performed using GATK Funcotator, incorporating gnomAD, EXAC, 1000 Genomes Project, ClinVar, and COSMIC databases, with functional impact scores from SIFT and PolyPhen. Variant data were used to assess frequency, pathogenicity, and distribution across study groups (AAs, NHWs, Ghanaians, and Ethiopians).

### Mutational landscape analysis

Annotated MAF files were imported into R using the MAFtools package (19). Variants were filtered at sequencing depth >20 and allele frequency <1% across population databases (AF_EXAC, gnomAD_exome_AF, AF_TGP all <0.01). To account for FFPE-associated cytosine deamination artifacts (C>T and G>A transitions), these mutations were subject to stricter thresholds: minimum depth of 50×, ≥8 variant-supporting reads, and VAF ≥10%, unless documented in COSMIC. TMB was calculated by dividing non-synonymous somatic mutations by the estimated coding genome size, with TMB-H defined as ≥10 mut/Mb and TMB-L as <10 mut/Mb. Mutational analyses focused on 11 genes implicated in key CRC pathways: *APC*, *BRAF*, *CTNNB1*, *FBXW7*, *KRAS*, *NRAS*, *PIK3CA*, *RNF43*, *SMAD4*, *TGFBR2*, and *TP53*.

### Comparing variants across populations

Variants in African-ancestry groups were compared to non-Hispanic Whites using a generalized linear model (GLM) with a binomial family. Statistical significance was set at p <0.05. Variant frequencies were estimated per population group to identify statistically significant differences across ancestral groups while controlling for sampling variation.

## RESULTS

We analyzed 150 CRC tumors from individuals undergoing care in the United States (43 AA and 53 NHW), West Africa (GHA, n = 21), and East Africa (ETH, n = 33) (**Table 1**). The ages at CRC diagnosis ranged from 16 to 78 years, with 64% of cases across all demographic groups classified as early-onset CRC. The proportion of early-onset cases in our cohort differed significantly by population group (*p*< 0.05), with more cases among individuals less than 50 years identified in the United States (92% NHW and 58% AA), than Africa (35% ETH and 43% GHA). The male-to-female ratios were also similar across all demographic groups (*p* > 0.05).

**Table 1:** Demographic, Clinical, and Molecular Characteristics of Colorectal Cancer Cases Across Population Groups. This table summarizes the demographic (age and sex), clinical and molecular features of colorectal cancer (CRC) cases across four population groups, African American (AA, n=43), Non-Hispanic White (NHW, n=53), Ghanaian (GHA, n=21), and Ethiopian (ETH, n=33). The clinical and molecular characteristics include primary tumor location, cancer stage, and key molecular markers such as *MLH1*/*PMS2*, MSH2/MSH6 mismatch repair (MMR) protein expression, *BRAF,* and *KRAS* status. Percentages are calculated based on the total number of samples tested within each group.

| Characteristic | AA<br>N = 43<sup>1</sup> | ETH<br>N = 33<sup>1</sup> | GHA<br>N = 21<sup>1</sup> | NHW<br>N = 53<sup>1</sup> | Pvalue |
| --- | --- | --- | --- | --- | --- |
| **Onset (Age at Diagnosis)** |  |  |  |  | **p < 0.001** |
| Early Onset (<50 years) | 25 (58%) | 11 (35%) | 9 (43%) | 49 (92%) |  |
| Late Onset (≥50 years) | 18 (42%) | 20 (65%) | 12 (57%) | 4 (8%) |  |
| **Sex** |  |  |  |  | **p = 0.7532** |
| Female | 21 (49%) | 13 (42%) | 12 (57%) | 27 (51%) |  |
| Male | 22 (51%) | 18 (58%) | 9 (43%) | 26 (49%) |  |
| **Primary Tumor Location** |  |  |  |  | **p < 0.001** |
| Any/ All Colon | 0 (0%) | 1 (3%) | 1 (5%) | 0 (0%) |  |
| Left Colon | 21 (50%) | 29 (94%) | 8 (40%) | 39 (75%) |  |
| Right Colon | 21 (50%) | 1 (3%) | 11 (55%) | 13 (25%) |  |
| **Stage** |  |  |  |  | *p* = 0.6252 |
| Stage 0 | 3 (7%) | 0 (0%) | 0 (0%) | 1 (2%) |  |
| Stage 1 | 9 (22%) | 1 (7%) | 4 (19%) | 9 (17%) |  |
| Stage 2 | 13 (32%) | 6 (43%) | 7 (34%) | 14 (27%) |  |
| Stage 3 | 12 (29%) | 7 (50%) | 10 (48%) | 22 (42%) |  |
| Stage 4 | 4 (10%) | 0 (0%) | 0 (0%) | 6 (12%) |  |
| **MMR Phenotype** |  |  |  |  |  |
| dMMR | 4 (25.0%) | 1 (3.0%) | 5 (31.3%) | 4 (9.3%) | *p* = 0.0132 |
| pMMR | 12 (75.0%) | 32 (97.0%) | 11 (68.8%) | 39 (90.7%) |  |
| **BRAF Status** |  |  |  |  | *p* = 0.0895 |
| Mutant | 0 (0%) | 0 (0%) | 2 (10%) | 2 (4%) |  |
| Wild Type | 42 (100%) | 33 (100%) | 19 (90%) | 49 (96%) |  |
| **KRAS Status** |  |  |  |  |  |
| Mutant | 18 (43%) | 15 (45%) | 1 (5%) | 19 (37%) | *p* = 0.0053 |
| Wild Type | 24 (57%) | 18 (55%) | 20 (95%) | 33 (63%) |  |

*N* = total samples available, *n* = samples tested, *P*value calculated using Fisher's Exact

### Clinical features of CRCs across demographic groups

There were no significant differences in tumor staging by population group (*p* > 0.05), with 40% of the CRC cases in our study were Stage 3 at diagnosis (29% of AA cases, 50% of Ethiopian cases, 48% of Ghanaian cases, and 42% of NHW cases; **Table 1**). We classified tumors as either right or left, with the latter including sigmoid and rectum tumors (**Table S2**). Right-sided tumors comprised 50% (21/42) among AAs, 55% (11/21) among Ghanaians, 25% (13/52) among NHWs, and 3% (1/31) among Ethiopians (*p* < 0.05). Rectal tumors were the most common among Ethiopians, comprising 68% (21/31), followed by NHW 35% (18/52), and were less common among AAs and Ghanaians, making up 14% (6/42) and 5% (1/21) of CRCs, respectively (**Table S2**). Sigmoid tumors were common amongst all the demographic groups, comprising 21% (9/42) AAs, 23% (7/31) Ethiopians, 30% (6/21) Ghanaians, and 29% (15/52) NHWs (**Table S2**).

### Standard clinical molecular profiling approaches reveal population-specific differences in CRC molecular features

There were significant differences in MMR status by population (*p* < 0.05), with 9% NHW, 15% AA, 33% Ghanaian, and 2% Ethiopian cases MMR deficient (dMMR). Mutation analyses showed that the dMMR phenotype was driven by *MLH1/PMS2* deficiency in 24% of AA cases, 3% of Ethiopian cases, 29% of Ghanaian cases, and 5% of NHW cases (**Table 1**), and *MSH2/MSH6* deficiency in 6% of AA cases, none (0%) of the Ethiopian cases, 21% of Ghanaian cases, and 5% of NHW cases. Individuals with deficient MMR (dMMR) were diagnosed at a lower mean age at diagnosis compared to those with proficient MMR (pMMR), although this difference was not statistically significant (**Figure S2**). The *BRAF* V600E mutation, commonly associated with poor prognosis and MSI-high tumors, was absent in AA and Ethiopian cases but was detected in 4% (2/53) of NHW tumors and 10% (2/21) of Ghanaian tumors (**Table 1**). For *KRAS* mutations, 43% of AA tumors carried mutations at codon 12/13, compared to 45% in Ethiopians, 37% in NHWs, and 5% in Ghanaians (*p* < 0.05) (**Table 1**).

### Tumor mutation burden distribution is significantly associated with population group

We utilized NGS DNA data to characterize the tumor mutation burden across the AA, Ghanaian, Ethiopian, and NHW CRC tumors (**Figure 1**). The sequencing quality of the tumor DNA was high across all the samples, assessed using sequencing depth (∼6 million reads per sample), GC content, and per-base quality scores (Phred scores). We classified samples as TMBH ≥10 mutations/Mb) or TMBL (<10 mutations/Mb) to evaluate the prevalence of hypermutation (**Figure 1**). We observed a statistically significant association between population and TMB classification (*p* < 0.05; **Figure 1**). Specifically, the Ethiopian population exhibited a distinct enrichment of TMBH (12/33, 36%) compared to the NHW (7/53, 13.2%), AA (4/43, 9.3%), and Ghanaian (0/21, 0%) populations. We did not observe significant associations between TMB status and MMR status, tumor stage, and primary tumor location. Stratification by biological sex revealed no significant difference in the proportion of TMB-High tumors between males and females within any population (**Figure 1**). Furthermore, TMB status was not significantly associated with anatomical site (Right vs. Left/Rectum; **Figure 1**), tumor stage (**Figure 1**), or *MLH1/PMS2* status (**Figure 1**). This lack of association with *MLH1/PMS2* status is particularly notable, suggesting that high mutational burden, specifically within the Ethiopian population, may be driven by mechanisms distinct from classical Mismatch Repair Deficiency (dMMR).

**Figure 1:**
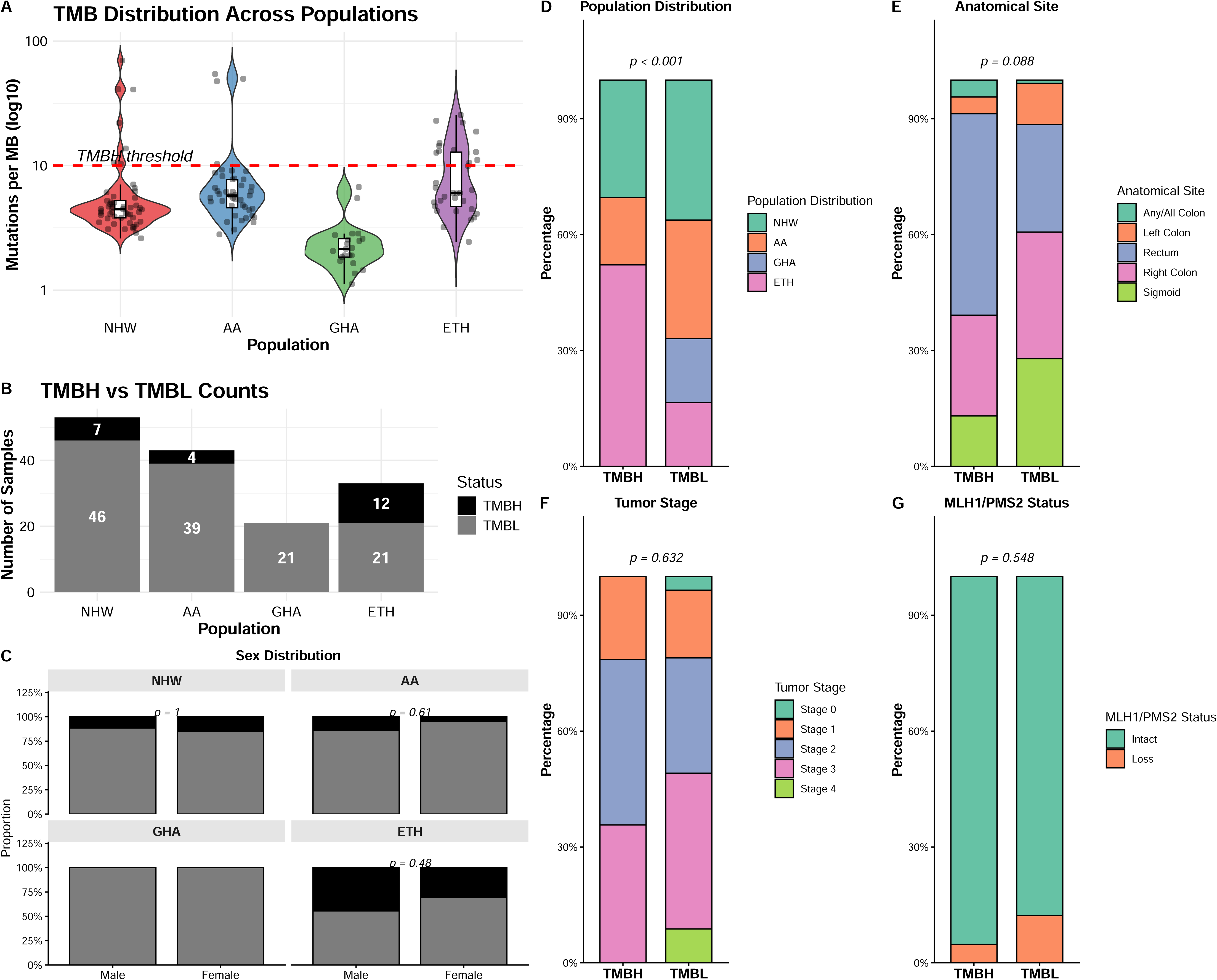
Distribution, stratification, and clinical determinants of Tumor Mutational Burden (TMB). (A) Distribution of mutations per megabase (log10 scale) across African American (AA), Ghanaian (GHA), Ethiopian (ETH), and Non-Hispanic White (NHW) cohorts. (B) Counts of TMB-High ≥10 mut/MB) versus TMB-Low samples (<10 mut/MB). (C–F) Stacked bar charts displaying the association between TMB status and (C) Sex, (D) Population, (E) Anatomical site, (F) Tumor Stage, and (F) MLH1/PMS2 Status. P-values indicate significance of sex-based differences within each group (Fisher’s Exact Test).

### Mutation rates are significantly different in key driver genes across populations

The somatic mutational landscape across the study populations revealed significant differences, suggesting that the molecular drivers of oncogenesis vary considerably by population group (**Table 2**). Analysis of the *Wnt* signaling pathway showed a striking divergence in how it is dysregulated across cohorts. While *APC* mutations were nearly ubiquitous in the AA (93.0%) and NHW (92.5%) cohorts, their prevalence was significantly lower in the Ethiopian (60.6%) and Ghanaian (38.1%) groups (*p* < 0.001). Interestingly, this deficit in *APC* alterations in the AA and Ghanaian cohorts appeared to be partially offset by a higher frequency of *CTNNB1* mutations (25.6% and 19.0%, respectively) compared to the NHW group, where such mutations were nearly absent at 1.9% (*p* = 0.0042). Similarly, *RNF43* mutations showed significant variation (*p* = 0.0047), reaching its highest mutational frequency in the NHW cohort. Specifically, *RNF43* was mutated in 73.6% of NHW samples, a rate nearly double that seen in the other populations. This shift indicates that while *Wnt* activation remains a central feature, the specific genetic hit favored by the tumor may be population-dependent. Further investigation into other key driver pathways highlighted additional disparities, where *PIK3CA* and *TP53* mutations showed significant variation (*p* = 0.0002 and *p* = 0.0057, respectively), with both genes reaching their highest mutational frequency in the AA and NHW cohorts, respectively. The AA cohort demonstrated a notably high *PIK3CA* mutational prevalence (39.5%) compared to the Ethiopian (3.0%) and Ghanaian (4.8%) cohorts, representing a highly significant distribution across the study groups (*p* = 0.0002). In contrast, mutations in *TGFβ* signaling components, such as *SMAD4* and *TGFBR2*, as well as *FBXW7* and *NRAS*, did not reach statistical significance, suggesting a more uniform, albeit lower, distribution across these specific genes regardless of cohort origin. These data collectively suggest that the genomic architecture of these tumors is not uniform across populations, which may influence disease course.

**Table 2:**
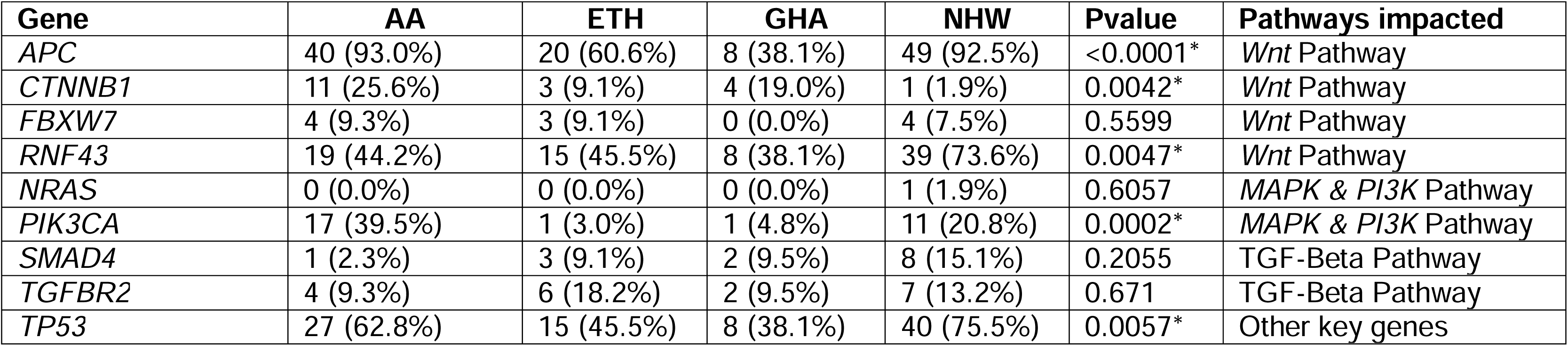
Somatic mutation rates in key colorectal cancer genes.

| Gene | AA | ETH | GHA | NHW | Pvalue | Pathways impacted |
| --- | --- | --- | --- | --- | --- | --- |
| APC | 40 (93.0%) | 20 (60.6%) | 8 (38.1%) | 49 (92.5%) | <0.0001* | Wnt Pathway |
| CTNNB1 | 11 (25.6%) | 3 (9.1%) | 4 (19.0%) | 1 (1.9%) | 0.0042* | Wnt Pathway |
| FBXW7 | 4 (9.3%) | 3 (9.1%) | 0 (0.0%) | 4 (7.5%) | 0.5599 | Wnt Pathway |
| RNF43 | 19 (44.2%) | 15 (45.5%) | 8 (38.1%) | 39 (73.6%) | 0.0047* | Wnt Pathway |
| NRAS | 0 (0.0%) | 0 (0.0%) | 0 (0.0%) | 1 (1.9%) | 0.6057 | MAPK & PI3K Pathway |
| PIK3CA | 17 (39.5%) | 1 (3.0%) | 1 (4.8%) | 11 (20.8%) | 0.0002* | MAPK & PI3K Pathway |
| SMAD4 | 1 (2.3%) | 3 (9.1%) | 2 (9.5%) | 8 (15.1%) | 0.2055 | TGF-Beta Pathway |
| TGFBR2 | 4 (9.3%) | 6 (18.2%) | 2 (9.5%) | 7 (13.2%) | 0.671 | TGF-Beta Pathway |
| TP53 | 27 (62.8%) | 15 (45.5%) | 8 (38.1%) | 40 (75.5%) | 0.0057* | Other key genes |

### Variants from American tumors are relatively well characterized compared to those from African populations

Next, we characterized the mutation landscape beyond simple counts by analyzing the clinical significance and predicted functional impact of somatic variants identified across populations, stratified by TMB status (**Figure 2**). We considered all the variants identified on the 290-gene Qiagen Comprehensive Cancer panel, and filtered out variants that had a clinical significance of benign. The majority of variants across all populations and TMB statuses were classified as “unknown” (grey bars), ranging from 87% in ETH TMBL to 92.4% in AA TMBH. The most “unknowns” were identified among AA and GHA tumors compared to NHW and ETH samples across TMBL and TMBH groups. The proportion of pathogenic variants was notably higher among AA (4.5% and 1.5%) and NHW (5.1% and 1.6%) across TMBL and TMBH tumors, respectively. This pattern was also observed for VUS among TMBH samples. In contrast, the proportion of pathogenic variants were 1.3% and 1.5% among Ethiopian and Ghanaian TMBL tumors respectively, and 0.8% among Ethiopian TMBH tumors.

**Figure 2:**
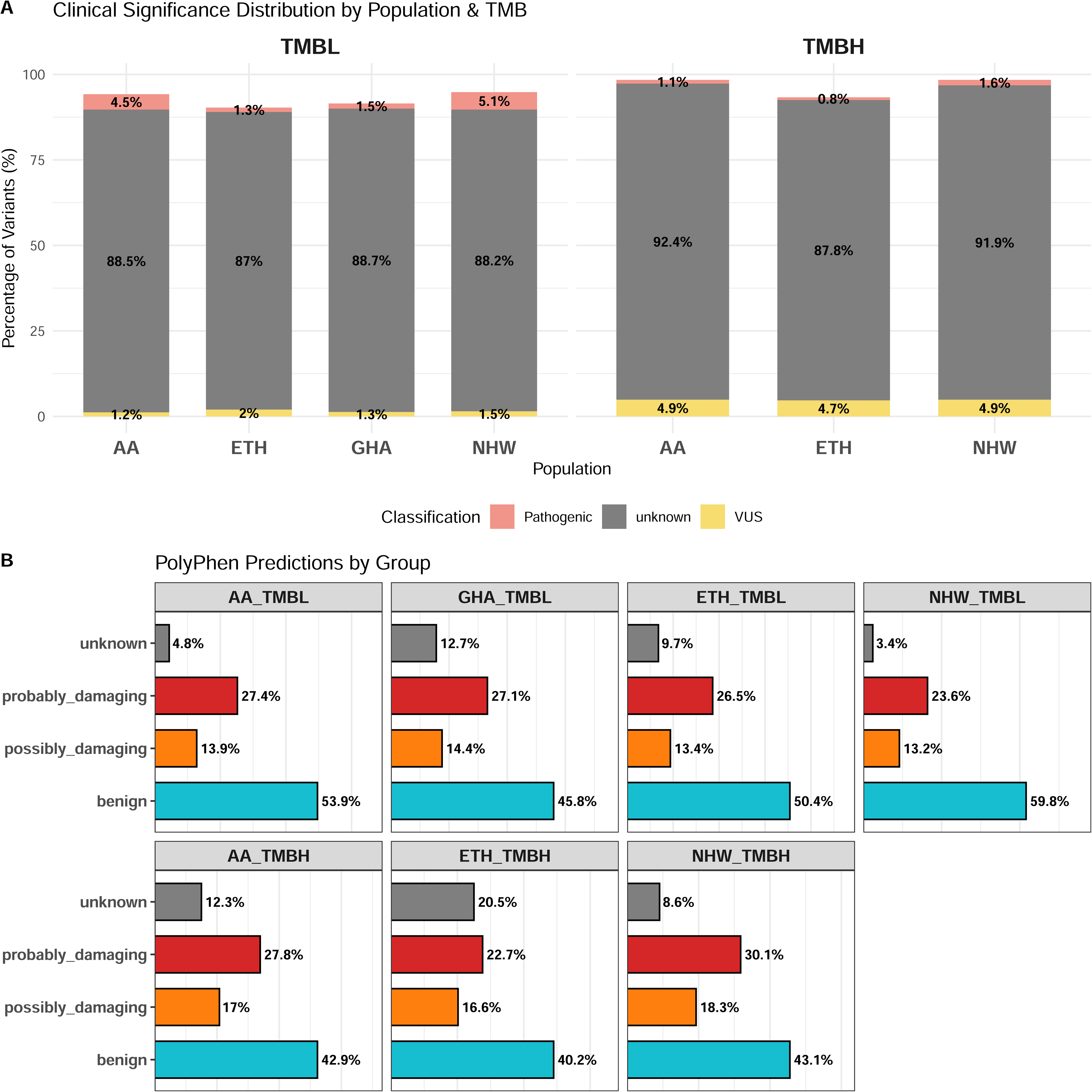
Clinical classification and computational functional assessment of somatic variants. (A) Stacked bar charts displaying the proportion of somatic variants classified by clinical significance (ClinVar) across African American (AA), Ghanaian (GHA), Ethiopian (ETH), and Non-Hispanic White (NHW) populations, stratified by TMB-High and TMB-Low status. (B) Functional impact predictions (PolyPhen) specifically for the subset of variants with “Unknown” clinical significance from Panel A, categorized as Benign, Possibly Damaging, or Probably Damaging.

Given the high prevalence of variants with unknown clinical significance, we utilized PolyPhen to predict functional impact (**Figure 2**). While the majority of ClinVar unknown variants were predicted to be benign (ranging from 40.2% to 59.8% across groups), a substantial fraction was predicted to be deleterious (possibly and probably damaging). In the TMBL group, variants predicted as “probably damaging” ranged between 26% to 28% among AA, Ghanaian, and Ethiopian populations, with the NHW population showing a slightly lower frequency (24%). Interestingly, TMBH tumors displayed a shift in functional distribution; compared to their TMBL counterparts, they exhibited a reduction in the proportion of benign variants and an increase in variants with “unknown”, “possibly damaging”, and “probably damaging” functional predictions. Higher proportions of “possibly/ probably damaging” variants were identified in AA and NHW TMBH tumors, as well as the lowest proportions of unknown variants. Seventy-four percent (74%) of the variants predicted to be probably damaging were absent in COSMIC (**Figure S3**), and their potential impact on the tumor biology of the different populations remains unexplored. Taken together, these results suggest either distinct tumor biology in individuals of African descent compared with those of European descent or that variants in tumors from individuals of European descent are better characterized than those in other populations.

### Tumors demonstrate population-specific recurrence of pathogenic and variants of uncertain significance across TMB strata

Analysis of pathogenic and VUS variant distribution across TMBH and TMBL CRC tumors revealed that the total number of recurrent variants was higher in TMBH tumors, consistent with the hypermutated phenotype of this stratum (**Figure S4**). Given the unbalanced distribution of TMBH tumors, concentrated predominantly in ETH (*n*=12) with no GHA representation, TMBH findings are presented as exploratory, and the primary focus of cross-population inference remains the TMBL cohort (**Figures S4 and S5**). In contrast to TMBL tumors, where 13 hotspot genes harboring greater than 10 variants were identifiable predominantly in AA and NHW populations (*PTPRJ*, *BRCA2*, *RAD50*, *POLE*, *BLM*, *BCL10*, *KRAS*, *BAX*, *MSH6*, *MSH3*, *AR*, *MED12*, and *BRCA1*), pathogenic and VUS variants in TMBH tumors were broadly distributed without discrete hotspot concentration. Within TMBH tumors, *CHEK2* Y224H and *POLE* V1446fs reached 100% and 75.0% recurrence respectively in ETH tumors, suggesting that hypermutation in Ethiopian CRC may be disproportionately driven by POLE proofreading defects and checkpoint dysregulation, though validation in larger East African cohorts is required.

In TMBL tumors, hierarchical clustering of variant recurrence profiles revealed that AA and NHW harbored the most similar mutational landscapes, while GHA exhibited a distinct pattern driven by population-enriched variants. ETH TMBL tumors showed negligible recurrent variant frequencies across all loci, likely reflecting the modest sample size, though potential true population-level differences cannot be excluded. Among pathogenic variants, convergent dysregulation of DNA damage response and homologous recombination was a shared feature of AA and NHW CRC, reflected by high-frequency recurrence of *POLE* V1446fs (AA: 74.4%; NHW: 54.3%), *BLM* N515fs (AA: 71.8%; NHW: 65.2%), and *BRCA2* I605fs (AA: 53.8%; NHW: 39.1%). GHA tumors were instead defined by markedly elevated *CHEK2* Y224H recurrence (61.9% versus 13.0% in NHW), alongside enrichment of *JAK3* E997* and *FGFR2* A86S, implicating population-specific cytokine and receptor tyrosine kinase pathway activation. In AA tumors, statistically significant enrichment of *CTNNB1* S45P implicated Wnt/β-catenin dysregulation as a distinguishing oncogenic feature relative to NHW.

### Ancestry-associated variant recurrence in TMBL colorectal cancer implicates population-specific oncogenic pathway dysregulation

Recurrent mismatch repair-associated VUS including *MSH3* K383fs (AA: 28.2%; NHW: 37.0%) and *MSH6* F1088fs (AA: 25.6%; NHW: 32.6%) were identified exclusively in AA and NHW TMBL tumors, suggesting these variants may contribute to genomic instability below the conventional TMB-high threshold with implications for immunotherapy eligibility in populations currently excluded from TMB-directed treatment pathways. Binomial GLM identified 20 of 33 recurrent TMBL variants as statistically significant (p<0.05) in at least one pairwise comparison against NHW (**Figure 3**, shown in bold). Cross-referencing TMBL and TMBH landscapes further identified a core set of 14 variants shared across both strata (**Figure S5**), suggesting these represent recurrent mutational events intrinsic to CRC biology rather than consequences of hypermutation alone, and candidates for functional evaluation in diverse population contexts.

**Figure 3:**
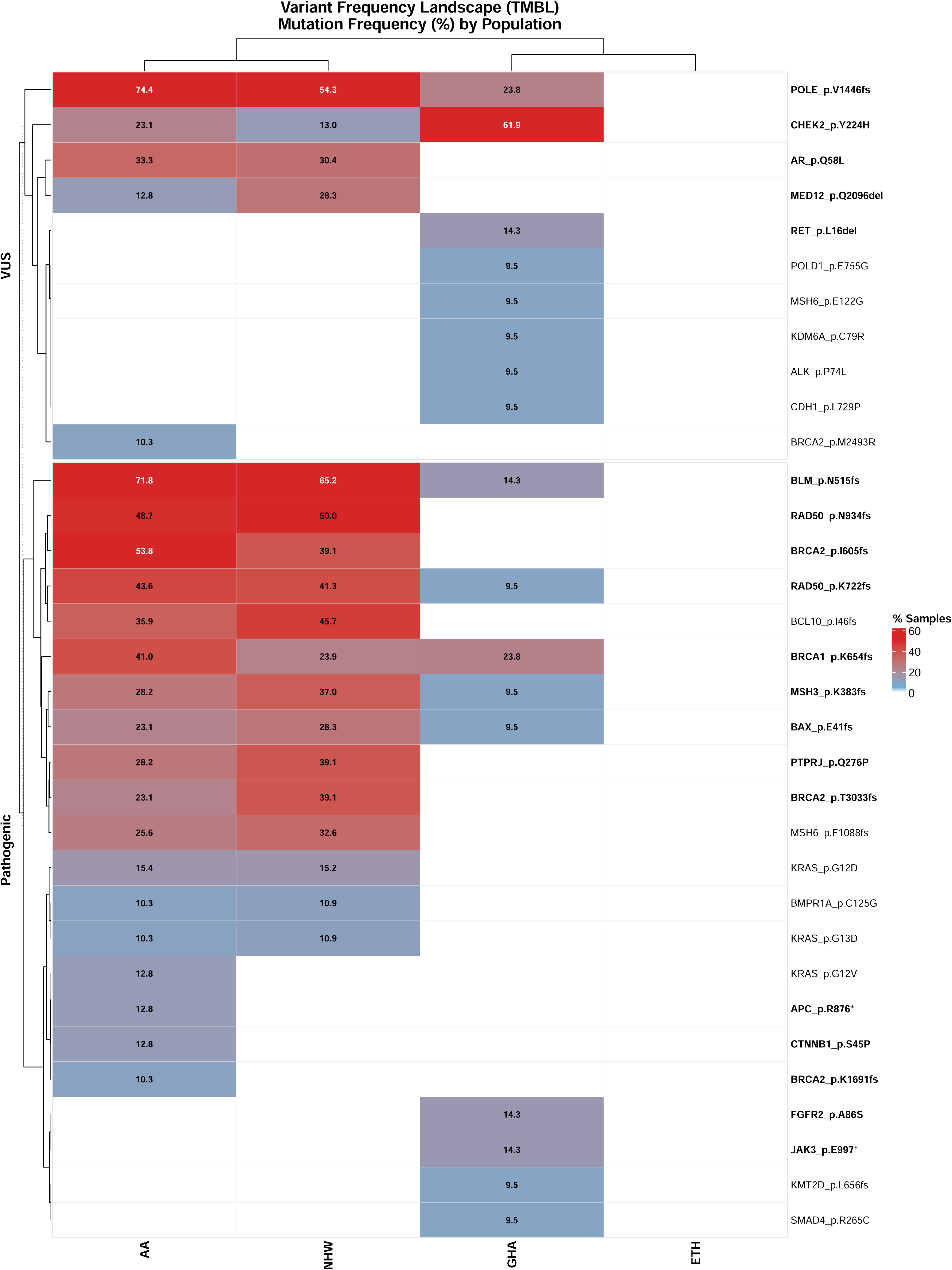
Frequency and recurrence of Pathogenic and VUS variants across diverse CRC populations. Heatmap displays the number of unique recurrent variants (found in >10% samples) classified as Pathogenic or Variant of Uncertain Significance (VUS). Panels are stratified by population (AA, NHW, GHA, ETH). The color gradient represents the recurrence frequency of each specific variant within that cohort: Blue indicates lower recurrence (10 −25%), pale red indicates intermediate recurrence (∼50%), and red indicates high recurrence (up to 100%). Specific gene and protein changes are labeled on the y-axis, and specific frequencies are shown in the panels. The asterisks (*) show the variants that are significantly different in one population (AA, GHA, or ETH) compared to the NHW population.

Together, these findings reveal clinically relevant, ancestry-associated mutational heterogeneity in TMBL CRC with direct implications for variant interpretation, risk stratification, and equitable therapeutic targeting in African and African-diaspora populations currently underrepresented in genomic databases and clinical trial cohorts.

## DISCUSSION

African Americans have the highest CRC mortality rate in the U.S, with approximately 18.5 deaths per 100,000 compared to 13.6 among non-Hispanic whites, and this disparity persists even after adjusting for access to care and socioeconomic factors, suggesting possible differences in tumor biology (20–25). Our study adds to the increasing evidence that biological differences in population groups may be a contributing factor to these disparities. We characterized the clinical and molecular features of CRC cases among African Americans, Ghanaians, Ethiopians, and non-Hispanic Whites, and we identified differences in mutation rates of key CRC driver genes and the occurrence of tumor somatic mutations by population. We also showed that variants in African populations are less characterized compared to those in tumors from the United States and identified mutational signatures that were common among US-derived tumors. These differences may influence disease progression pathways, providing insights into population-specific tumor biology. This also highlights the need for studies in populations of African descent to characterize the significance of previously unrecognized variants.

Databases, such as COSMIC and TCGA, utilized samples obtained from studies that included mostly NHWs individuals, with very few individuals of African ancestry (5). The TCGA data contains 500 African American cases out of a total of 5899, making up only 8.5% of the cohort. Our studies and others that utilize samples from diverse populations may continue to uncover potentially novel variants important to CRC progression. Several of the somatic variants that we identified differed in occurrence by population group, were predicted to have deleterious and damaging consequences and have also not been reported in the literature.

When we investigated common CRC mutations, such as the *BRAF* V600E variant, which has been reported to occur in approximately 8-12% of all CRC cases, this specific variant was predictably rare across all the populations we studied. However, other non-*V600E* somatic variants were found in up to 43% and 44% of African American and non-Hispanic white CRCs, respectively, while mutation rates of this gene were low among Ghanaian and Ethiopian samples (14% and 33%, respectively). The *BRAF* gene, which encodes a serine/threonine kinase, is involved in the *MAPK/ERK* signaling pathway that is critical for regulating cell growth, cell division, differentiation, and cell survival, and mutations in this gene are associated with more aggressive disease with poorer prognosis. Our results might differ from previous studies as our population group was enriched for early-onset cases, unlike previous studies that included mostly older patients. Similarly, we identified high somatic mutation rates in the *KRAS* and *APC* genes of AAd and NHWs compared to African samples. This is also corroborated in a previous study that also reported fewer *APC* mutations and *WNT* pathway alterations from African samples collected in Nigeria (16). These findings suggest differences in CRC subtype distribution by genetic ancestry, or by geography, probably influenced by environmental or lifestyle factors specific to location.

The results from this study have implications for how CRC is diagnosed and treated based on the different mutational profiles observed by population group. Many current therapeutic strategies and biomarkers, such as the *BRAF V600E* mutation and *KRAS* codon 12 mutations, are based on datasets that predominantly include European ancestry populations, potentially limiting their applicability to African ancestry populations and other underrepresented groups (5). Differences in these molecular profiles influence disease course, emphasizing the limitations of applying generalized CRC molecular classifications across diverse populations without considering ancestry-specific genomic variations. These findings reinforce the necessity of expanding genomic studies to underrepresented populations to capture the biological diversity of CRC and enhance personalized treatment strategies.

Our dataset was unique in that it included CRC cases from diverse ancestral backgrounds from the United States (AAs and NHWs), Ghana, and Ethiopia. This broad representation allowed for a comparative analysis of population-specific molecular alterations, shedding light on ancestry-related differences in tumor biology. Notably, by design, 64% of the cases were classified as early-onset CRC, diagnosed before the age of 50, which is significantly higher than the proportion typically observed in the TCGA genomic dataset for colorectal cancer (1). This high prevalence of early-onset cases in our dataset highlights the growing incidence of CRC among young individuals, regardless of population group, emphasizing the urgent need to understand genetic, environmental, and lifestyle factors contributing to this trend. By incorporating diverse population groups, our study provides valuable insights into the molecular landscape of CRC across different ethnic backgrounds, which may have important implications for risk stratification, targeted therapy development, and personalized treatment approaches.

Our study, however, was limited in that the multigene panel used for somatic mutation profiling analyzed only 290 genes, representing a subset of the total genes in the genome. As a result, additional oncogenic mutations, structural variations, and non-coding regulatory alterations may not have been captured. Future studies can be improved by expanding the genomic regions analyzed through whole-exome sequencing (WES) or whole-genome sequencing (WGS), which would allow for a more comprehensive assessment of somatic and germline variations contributing to colorectal cancer across diverse populations, including performing genetic ancestry testing *in lieu* of self-reported race. In addition, our analysis was based on a relatively small sample size, and the tumors included were convenience samples, which may limit the generalizability if the findings. Prognostic data were missing for many study subjects, preventing correlation of the somatic mutations with clinical outcomes. Finally, somatic DNA sequencing was performed without matched germline DNA, raising the possibility that some tumor variants identified may reflect benign germline polymorphisms rather than true somatic mutations. Addressing these limitations in future studies will be essential to advancing our understanding of colorectal cancer biology, particularly in underrepresented populations.

Despite these limitations, the insights from this study provide a valuable foundation for larger-scale genomic investigations. Population-specific genetic variants have been found to play a significant role in cancer aggressiveness in other cancers (11,12). By identifying population-specific mutational profiles in CRC, this research underscores the importance of including diverse ancestry groups in cancer genomics studies to improve risk stratification, biomarker discovery, and personalized treatment approaches. Expanding these findings in future studies could enhance our understanding of CRC pathogenesis and contribute to the development of precision medicine strategies tailored to underrepresented populations.

## CONCLUSIONS

This study provides a comprehensive characterization of somatic mutational profiles in CRC tumors from early- and late-onset cases across AAs, Ghanaians, Ethiopians, and non-Hispanic Whites NHWs. Although some population-specific differences were observed, the overall mutational landscape appeared to be more strongly influenced by geographic context and age of onset, with AA tumors exhibiting patterns similar to those observed in NHW tumors, both of which consisted predominantly of early-onset cases. These findings suggest that age- and geography-related factors, including environmental exposures and lifestyle influences, may contribute to shaping CRC tumor phenotypes. Expanding genomic studies to include globally diverse populations and age groups will be essential for improving our understanding of CRC biology and for informing more equitable precision oncology strategies.

## Acknowledgements

The authors thank the clinical pathologists at Georgetown University and Komfo Anokye Teaching Hospital for their contributions to tumor slide review and the University of Michigan Genomics Core for sequencing support. We are grateful to the patients and institutions in Ghana, Ethiopia, and the United States who contributed samples to this study. We also thank the Georgetown Lombardi Comprehensive Cancer Center for its support. This work was supported by the National Institutes of Health, National Cancer Institute (NIH/NCI R01-CA259420; LR, ES, KP, BM) and the American Cancer Society (IRG-23-1156148-27-IRG; BM).

## Data and code availability

The data that support the findings of this study contain protected patient information and cannot be publicly released. Researchers interested in accessing the data may submit a reasonable request to the corresponding author. Access will be considered on a case-by-case basis and is subject to institutional review board (IRB) approval and the execution of a data use agreement to ensure patient confidentiality and compliance with applicable regulations.

## SUPPLEMENTARY INFORMATION

### SUPPLEMENTARY TABLES

**Table S1:** Genes included in the Qiagen Comprehensive Cancer Panel.

**Table S2:** Distribution of primary tumor location across four population groups.

### SUPPLEMENTARY FIGURES

**Figure S1**: Computational pipeline for somatic variant detection and population-level mutational analysis.

**Figure S2**: Age at diagnosis distribution by Mismatch Repair (MMR) status across diverse CRC populations.

**Figure S3**: Frequency and recurrence of variants predicted as “Probably Damaging” in PolyPhen. Heatmap displays the number of unique recurrent variants (found in >10% samples & greater than 2 samples) classified as Absent or Present in the COSMIC database. Panels are stratified by population (AA, NHW, GHA, ETH). The color gradient represents the recurrence frequency of each specific variant within that cohort: Blue indicates lower recurrence (10 −25%), pale red indicates intermediate recurrence (∼50%), and red indicates high recurrence (up to 100%). Specific gene and protein changes are labeled on the y-axis, and specific frequencies are shown in the panels.

**Figure S4**: Distribution of Pathogenic and VUS Variants Stratified by Tumor Mutation Burden and Population Group.

**Figure S5**: Frequency and recurrence of Pathogenic and VUS variants across diverse CRC populations. Heatmap displays the number of unique recurrent variants (found in >10% samples) classified as Pathogenic or Variant of Uncertain Significance (VUS). Panels are stratified by population (AA, NHW, GHA, ETH). The color gradient represents the recurrence frequency of each specific variant within that cohort: Blue indicates lower recurrence (10 −25%), pale red indicates intermediate recurrence (∼50%), and red indicates high recurrence (up to 100%). Specific gene and protein changes are labeled on the y-axis, and specific frequencies are shown in the panels.

