## Supplementary Information for "Somatic Mutation Profiles in Colorectal Cancers Differ by Population"

**Figure S1**: Computational pipeline for somatic variant detection and population-level mutational analysis.

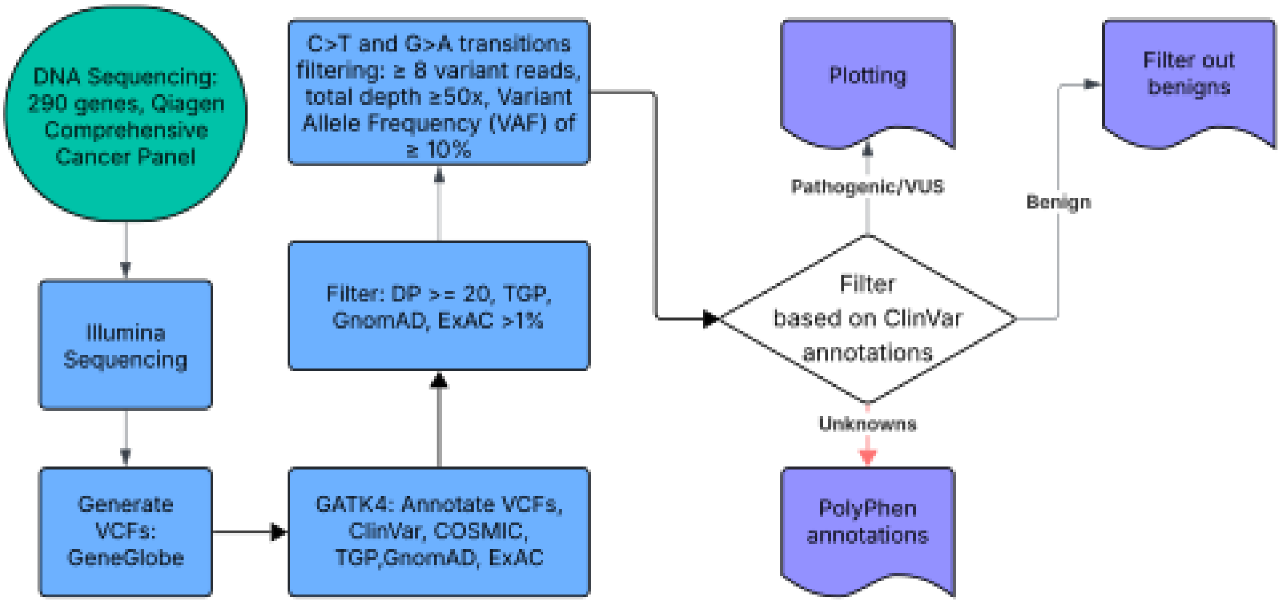

**Figure S2**: Age at diagnosis distribution by Mismatch Repair (MMR) status across diverse CRC populations.

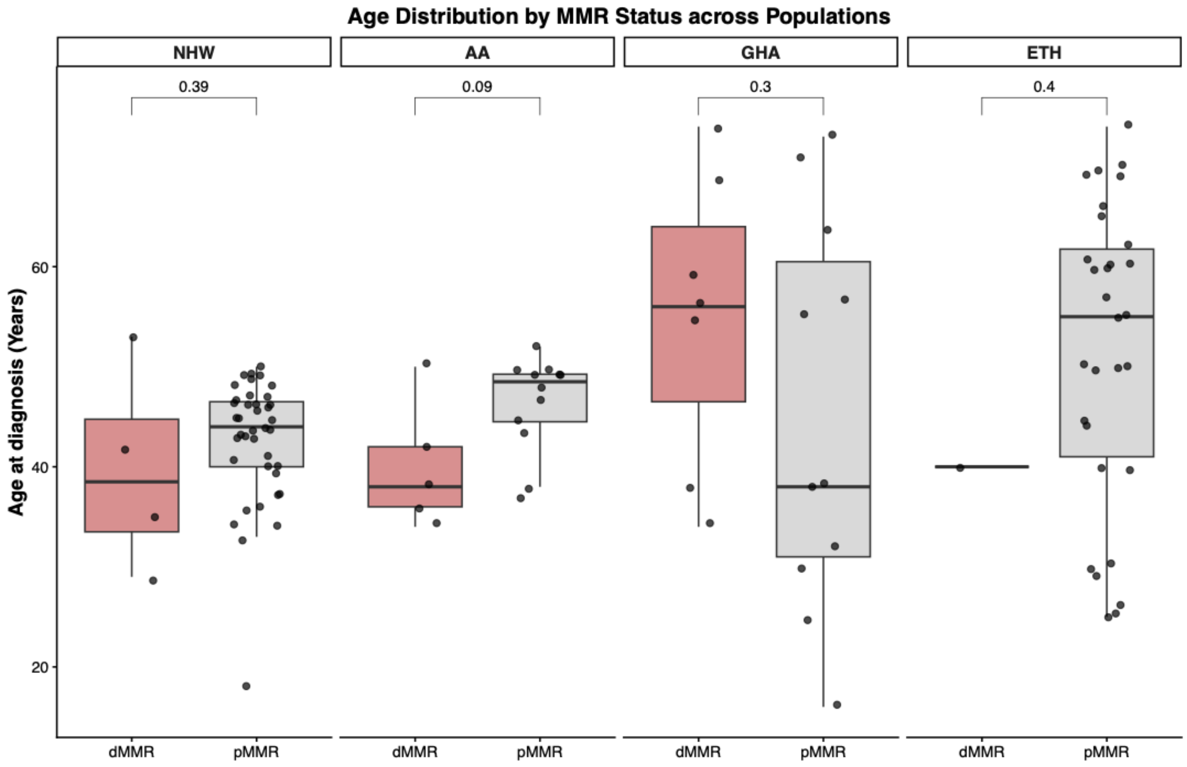

**Figure S3**: Frequency and recurrence of variants predicted as “Probably Damaging” in PolyPhen. Heatmap displays the number of unique recurrent variants (found in >10% samples & greater than 2 samples) classified as Absent or Present in the COSMIC database. Panels are stratified by population (AA, NHW, GHA, ETH). The color gradient represents the recurrence frequency of each specific variant within that cohort: Blue indicates lower recurrence (10 -25%), pale red indicates intermediate recurrence (~50%), and red indicates high recurrence (up to 100%). Specific gene and protein changes are labeled on the y-axis, and specific frequencies are shown in the panels.

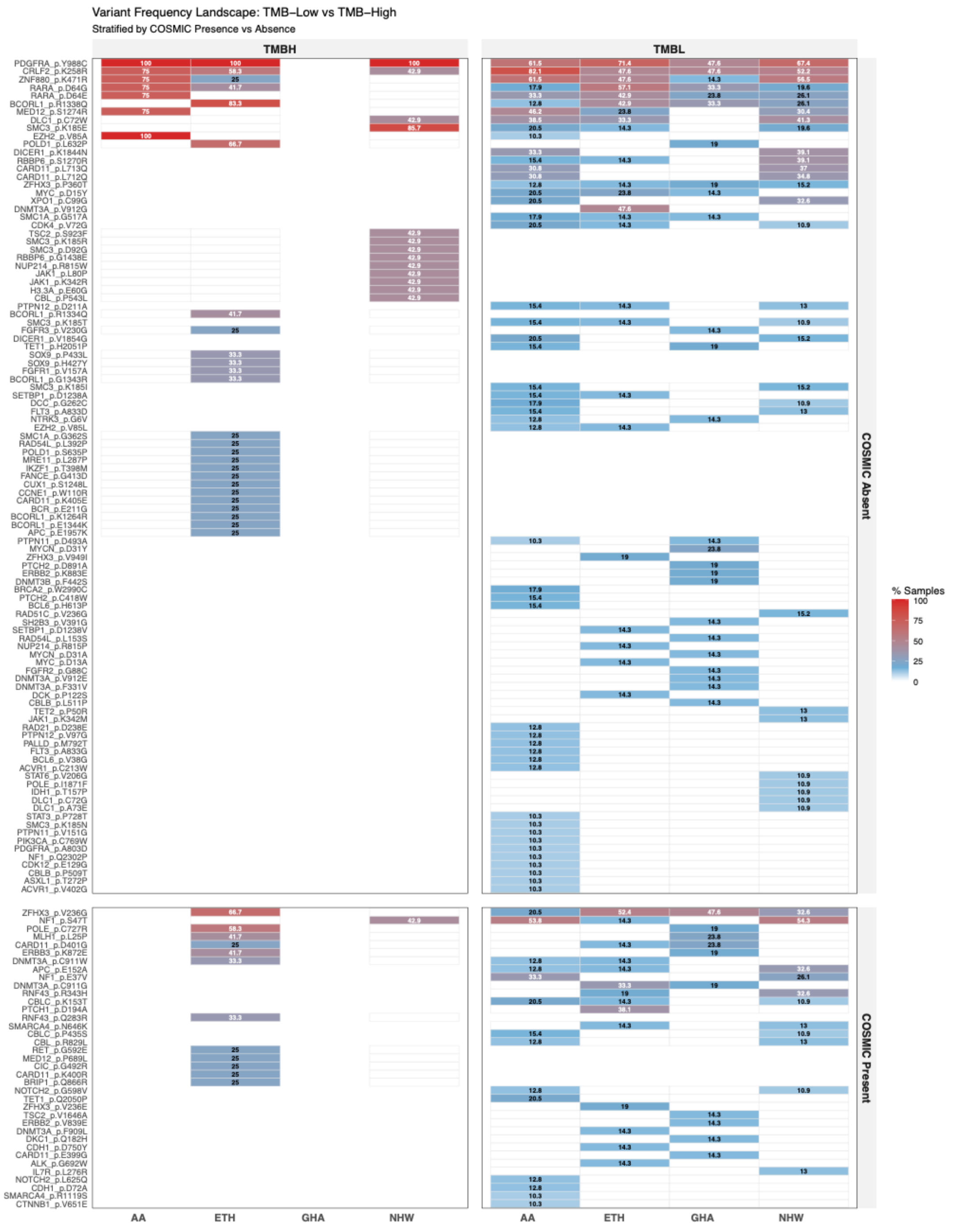

**Figure S4**: Distribution of Pathogenic and VUS Variants Stratified by Tumor Mutation Burden and Population Group.

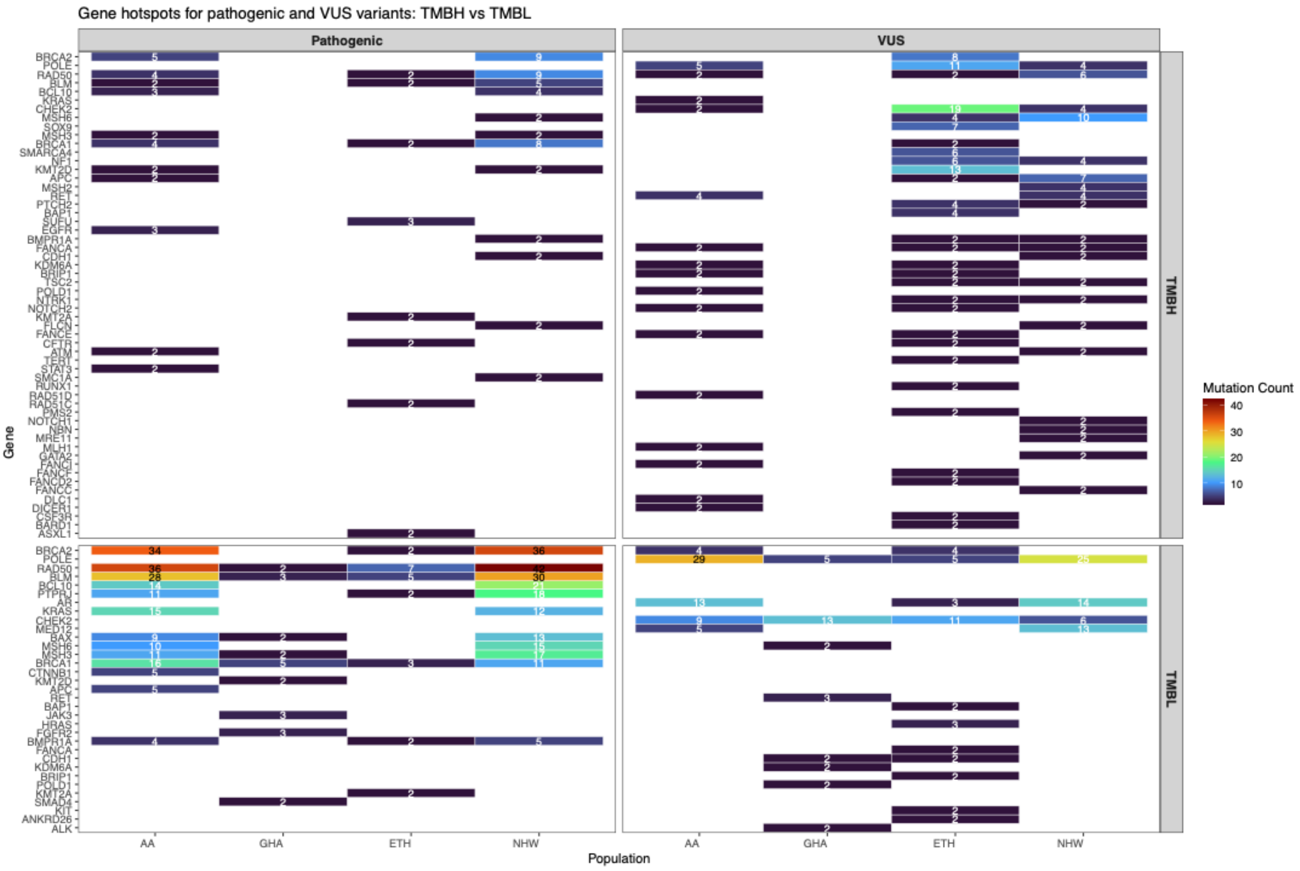

**Figure S5**: Frequency and recurrence of Pathogenic and VUS variants across diverse CRC populations. Heatmap displays the number of unique recurrent variants (found in >10% samples) classified as Pathogenic or Variant of Uncertain Significance (VUS). Panels are stratified by population (AA, NHW, GHA, ETH). The color gradient represents the recurrence frequency of each specific variant within that cohort: Blue indicates lower recurrence (10 -25%), pale red indicates intermediate recurrence (~50%), and red indicates high recurrence (up to 100%). Specific gene and protein changes are labeled on the y-axis, and specific frequencies are shown in the panels.

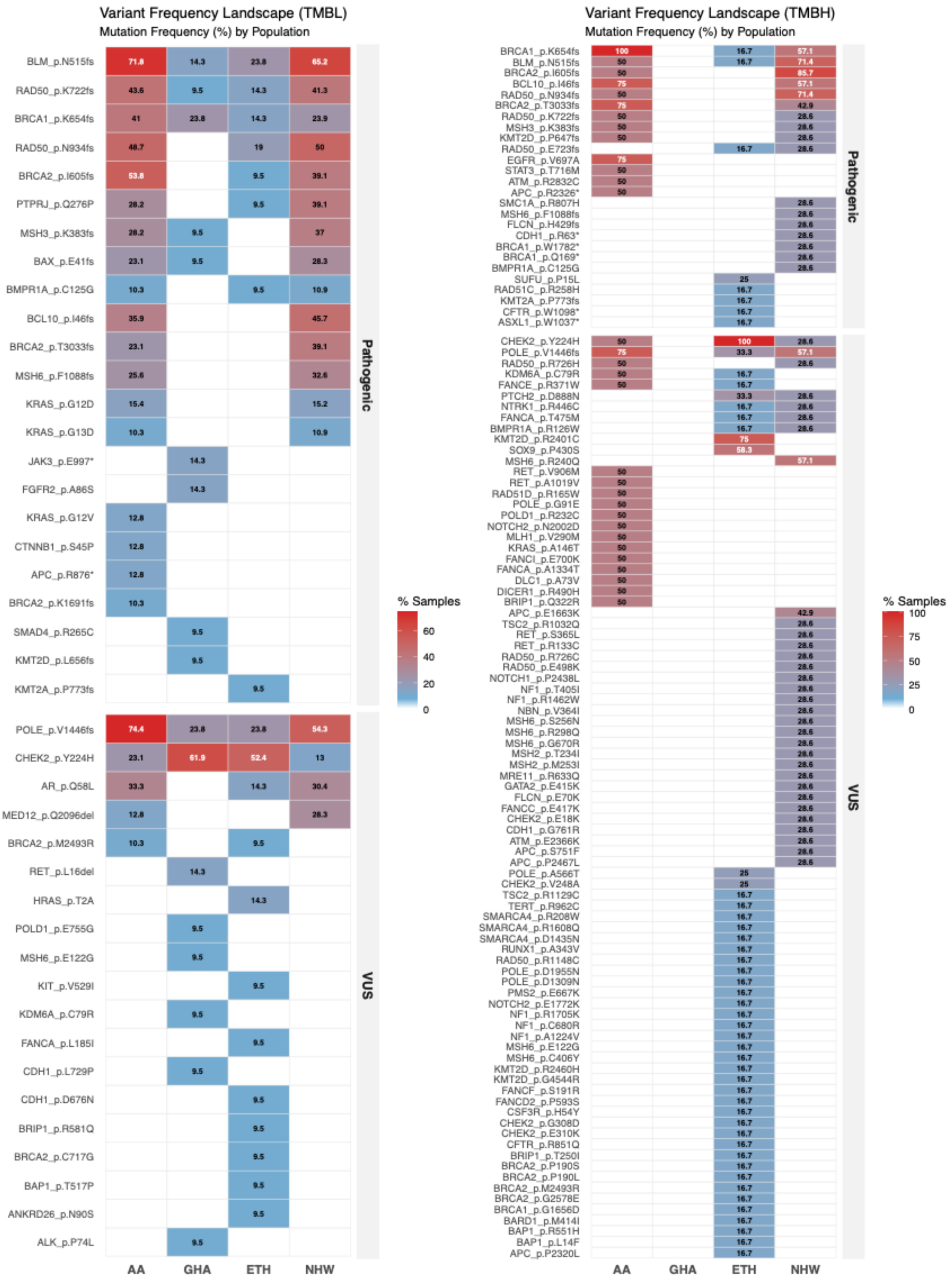

**Table S1**: Genes included in the Qiagen Comprehensive Cancer Panel.

| | | | | | | |
|---|---|---|---|---|---|---|
| *GNAS* | *RARA* | *SAMD9* | *EBF1* | *SETD2* | *CDC25C* | *RASGRP4* |
| *SETBP1* | *RUNX1* | *SAMD9L* | *DDX41* | *BAP1* | *ARHGAP26* | *CIC* |
| *NF1* | *ERG* | *CUX1* | *H1-4* | *ROBO2* | *CREBBP* | *ERBB3* |
| *CDK12* | *BCR* | *MET* | *FANCE* | *FOXL2* | *RBBP6* | *POLD1* |
| *TP53* | *CHEK2* | *CFTR* | *MSH2* | *ATR* | *ALK* | *PPP2R1A* |
| *STAT5B* | *EP300* | *BRAF* | *MSH6* | *BCL6* | *EPCAM* | *U2AF2* |
| *BRCA1* | *CYP2D6* | *DLC1* | *ZAP70* | *FGFR3* | *ROS1* | *MYCN* |
| *ETV4* | *ASXL1* | *PDGFRL* | *GLI2* | *CD38* | *TNFAIP3* | *GEN1* |
| *RNF43* | *EGFR* | *CSF3R* | *CXCR4* | *PDGFRA* | *ARID1B* | *DNMT3A* |
| *GNA13* | *PTPN12* | *MPL* | *PMS1* | *KIT* | *PRKN* | *KMT2A* |
| *AXIN2* | *GNB1* | *PTCH2* | *CARD11* | *DCK* | *MAD1L1* | *CBLC* |
| *SOX9* | *TNFRSF14* | *PIK3R3* | *PMS2* | *ABRAXAS1* | *NOTCH2* | *BAX* |
| *SRSF2* | *CDH1* | *RAD54L* | *KMT2D* | *TET2* | *DDR2* | *CDKN2A* |
| *FANCA* | *ZFHX3* | *JAK1* | *FGFR1* | *TERT* | *ABL2* | *CDKN2B* |
| *BRIP1* | *SF3B1* | *BCL10* | *RB1CC1* | *IL7R* | *GATA3* | *PAX5* |
| *PRPF8* | *IKZF2* | *DPYD* | *PREX2* | *MSH3* | *MAP3K8* | *PTCH1* |
| *DNMT3B* | *UGT1A1* | *BCL2* | *NBN* | *APC* | *JAK3* | *ABL1* |
| *SRC* | *ETV1* | *STK11* | *RAD21* | *MCC* | *MEF2B* | *RET* |
| *MTOR* | *IKZF1* | *SMARCA4* | *FANCD2* | *IRF1* | *CCNE1* | *TET1* |
| *ARID1A* | *CALR* | *PDGFRB* | *TGFBR2* | *RAD50* | *CEBPA* | *MXI1* |
| *SMC3* | *PTPN11* | *MAP2K1* | *MRE11* | *FBXW7* | *CDK6* | *FUBP1* |
| *TCF7L2* | *POLE* | *DICER1* | *CCND2* | *ZRSR2* | *LUC7L2* | *NRAS* |
| *CBL* | *BRCA2* | *BUB1B* | *BTK* | *CD79A* | *FGFR2* | *SMAD4* |
| *OPCML* | *FANCM* | *TSC2* | *BCOR* | *ERCC1* | *AKT1* | *PRSS1* |
| *CDKN1B* | *RAD51B* | *BLM* | *KDM6A* | *RAF1* | *RAD52* | *ETV6* |
| *ETNK1* | *MLH3* | *RAD51C* | *WT1* | *MLH1* | *FLCN* | *RB1* |
| *NUMA1* | *GATA1* | *CHEK1* | *EPHB2* | *CBLB* | *BMPR1A* | *FOXO1* |
| *MYC* | *SMC1A* | *DCC* | *IL23R* | *SMARCB1* | *ZEB2* | *H3C2* |
| *JAK2* | *AR* | *STAT3* | *NTRK1* | *ERBB2* | *ACVR1* | *SPOP* |
| *ATM* | *MED12* | *CRLF2* | *SUFU* | *RPS14* | *CTNNB1* | *MYD88* |
| *PPP2R1B* | *ATRX* | *CASP10* | *ZNF880* | *PIM1* | *H3-3A* | *PICALM* |
| *NUP214* | *STAG2* | *IDH1* | *XPO1* | *ELANE* | *KLF6* | *KRAS* |
| *NOTCH1* | *IDH2* | *HRAS* | *BUB1* | *PTEN* | *FANCC* | *CCND1* |
| *SLC22A18* | *STAT6* | *ETV5* | *PALB2* | *PIK3CA* | *NF2* | *INSRR* |
| *FANCF* | *DKC1* | *DHX15* | *B2M* | *RHOA* | *BIRC3* | *TYMS* |
| *PTPRJ* | *FOXA1* | *PALLD* | *RAD54B* | *GATA2* | *PPM1D* | *KLF2* |
| *BCORL1* | *NTRK3* | *PIK3R1* | *PLCG2* | *FANCG* | *CD274* | *FANCL* |
| *CDK4* | *FANCI* | *ANKRD26* | *RPA1* | *CBFB* | *ESR1* | *HSPH1* |
| *MDM2* | *FLT3* | *BARD1* | *EZH2* | *IKZF3* | *PHF6* | *RAD51D* |
| *SH2B3* | *INO80* | *SLC29A1* | *SRP72* | *PPP2R2A* | *CCR4* | *SOCS1* |
| *MUTYH* | *NPM1* | *PIGA* | *TOE1* | *SPINK1* | *FSBP* | *CD79B* |
| *DHX8* | *RAD51* | *UGT1A6* | | | | |

**Table S2**: Distribution of primary tumor location across four population groups.

| Characteristic | AA N = 43[1] | ETH N = 33[1] | GHA N = 21[1] | NHW N = 53[1] |
|---|---|---|---|---|
| **Primary Tumor Location** | | | | |
| Any/All Colon | 0 (0%) | 1 (3%) | 1 (5%) | 0 (0%) |
| Left Colon | 6 (14%) | 1 (3%) | 1 (5%) | 6 (12%) |
| Rectum | 6 (14%) | 21 (68%) | 1 (5%) | 18 (35%) |
| Right Colon | 21 (50%) | 1 (3%) | 11 (55%) | 13 (25%) |
| Sigmoid | 9 (21%) | 7 (23%) | 6 (30%) | 15 (29%) |

[1] n (%)

*N = total samples available, n = samples tested*
